# Performance of digital Early Warning Score (NEWS2) in a cardiac specialist setting: retrospective cohort study

**DOI:** 10.1101/2022.06.09.22275676

**Authors:** Baneen Alhmoud, Timothy Bonnici, Daniel Melley, Riyaz Patel, Amitava Banerjee

**Author notes:** Corresponding author: Baneen Alhmoud, Institute of Health Informatics, 222 Euston Road, London. NW1 2DA.

## Abstract

**Introduction:** Patients with Cardiovascular diseases (CVD) are at significant risk of developing critical events. Early warning scores are recommended for early recognition and rapid response to deteriorating patients, yet their performance has been poorly studied in cardiac care settings. Standardisation and integrated National early warning score (NEWS2) in EHRs are recommended but their evaluation in specialist settings is limited.

**Objective:** To investigate the performance of digital NEWS2 in predicting critical events: death, ICU admission, Cardiac arrest, and medical emergencies.

**Methods:** Retrospective cohort analysis

**Study cohort:** Individuals admitted with cardiovascular disease diagnoses in 2020 and patients with positive COVID-19.

**Measures:** We tested the ability of NEWS2 in predicting death, ICU admission, cardiac arrest, and medical emergency from admission and within 24 hours before the event. NEWS2 was supplemented with age and cardiac rhythm and investigated. We used logistic regression analysis with the area under the receiver operating curve (AUC) to measure discrimination.

**Results:** In 6143 patients admitted under cardiac speciality, NEWS2 showed moderate to low predictive accuracy (AUC: 0.63, 0.56, 0.7& 0.63; 95% CI). Supplemented NEWS2 with age showed no improvement while age and cardiac rhythm improved discrimination (AUC: 0.75, 0.84, 0.95 & 0.94; 95%CI). Improved Performance was found of NEWS2 for COVID-19 cases with age (AUC: 0.96, 0.7, 0.87& 0.88; 95% CI).

**Conclusion:** The performance of NEWS2 in patients with CVD is suboptimal, and fair for patients with COVID-19 to predict deterioration early. Adjustment with variables that strongly correlate with critical cardiovascular outcomes, i.e. cardiac rhythm, can improve the early scoring models. There is a need to define critical endpoints, engagement with clinical experts in development of models and further validation and implementation studies of EHR-integrated EWS in cardiac specialist settings.

## Introduction

Disease severity classification of patients with cardiovascular diseases (CVD) is challenging for nurses and physicians. Individuals with CVD can present with various sorts of critical events due to the disease’s pathophysiology or the comorbidities associated (1). The aetiology of the disease and the specialised care provided may impose the standardised use of deterioration risk scores, such as the widely adopted National Early Warning Score 2 (NEWS2) (2). Heterogeneity in the process of patient deterioration complicates detection and escalation.

CVD are the leading cause of death in the UK and worldwide, with an estimated health care cost of £9 billion annually (1,3). Short-term critical events, such as cardiac arrest and transfer to the Intensive Care Unit (ICU), are common in patients with CVD (4,5). In addition, mortality and morbidity are major concerns in patients with CVD globally (1). Risk stratification tools for long term outcomes have long been favoured in this disease subgroup. Models like the Global Registry of Acute Coronary Events (GRACE) and the CHADS2-VASc are validated long term risk scores in cardiovascular diseases (6,7). For early stage risk prediction, risk stratification of critical deterioration was unified for all disease groups and settings using early warning scoring systems (EWS) (8,9).

The recently implemented and developed NEWS2 has been recommended by the consensus of clinical experts. A minimum set of physiological parameters and standardised application across the NHS to promote patient safety and unify clinicians’ practice were the aims of NEWS2 (9–11). The predictability of NEWS and NEWS2 of critical events was found fair to acceptable to the emergency department (12,13) and medical and surgical settings. On the other hand, issues reported in specialised settings, like poor predictive performance in haematology settings (13) and the need to supplement NEWS in emergency settings (12,14) and for Covid patients (9), indicate its inefficiency in particular contexts. In cardiac settings, studies on EWS performance in critical events are poor and insufficient to defend the application for escalating the acutely ill. A Study in 2012 showed high predictive ability in cardiac patients using ViEWS, a model with different parameters than NEWS (15). Another examined RACE, a postoperative cardiac EWS tool for cardiac surgical ICU; a narrowly included subset of cardiac patients (16). Little is known about the predictive value of some NEWS2 components to be deemed reliable in this subgroup, i.e. inclusion of temperature or missing heart rhythm. Despite the need for reliable early deterioration detection, it is reported that specialised cardiac centres may overlook the value of developing and validating EWS. The complexity of models and difficulties in analysing electronic health records (EHR) data formed barriers to the validation of EWS (16). However, in the era of predictive modelling generated from EHRs, and EWS embedded in EHR, validating EWS in patients population with a high rate of critical events is necessary.

## Objective

To investigate the performance of digital NEWS2 in predicting critical events, at admission and prior to deterioration, for cardiac patients in the COVID-19 context in a cardiac specialist setting.

### Our specific aims are

1. To explore the independent association of physiological parameters and NEWS2 at hospital admission and 24hr prior critical events; with disease severity (ICU admission, Cardiac arrest, Medical emergency and death);
2. To examine the predictive ability of NEWS2 and the supplemented NEWS2 with potential determinants of disease severity on admission and 24 hrs prior to the condition worsening.
3. To compare the predictive value of NEWS2 with supplemented NEWS2 models.

## Methods

### Ethics

This study is registered and approved by the Health Research Authority (HRA) and Health and Care Research Wales (HCRW) and sponsored by University College London (UCL). REC reference: 20/PR/0286. A confirmation of capacity has been received from St Bartholomew’s hospital.

### Study cohort

The study population was defined as adult patients admitted to St Bartholomew’s hospital, a cardiac specialist and teaching hospital in London with heart and cancer centres, from January to December 2020, under cardiac speciality care; for more than 24 hours. Due to the nature of the pandemic, we have identified patients with Covid-19 based on positive PCR test results upon or during admission.

### Measures

#### NEWS2 and physiological parameters

We included physiological parameters and NEWS2 scores routinely obtained at hospital admission and 24 hours prior to deterioration. Included parameters that form NEWS2 score, were respiratory rate (breaths per minute), oxygen saturation (%), systolic blood pressure (mmHg), heart rate (beats/min), temperature (°C), and consciousness (measured by Glasgow Coma Scale (GCS) total score. We also included diastolic blood pressure, which is not part of NEWS2. Measurements time was chosen as the most completed set of parameters; measurements done 48 hrs and 7 days prior to events were not included due to missing and inconsistent data. Heart rhythm was included 24 hrs prior to event due to measurements recording by Cardiac Resuscitation Team (CRT).

#### Outcomes

The primary outcome was patients’ critical status following assessment on admission or 24 hours prior to a critical event. Outcomes were critical events categorised as in-hospital death, transfer to ICU, developing cardiac arrest, and medical emergency. A medical emergency was defined as deterioration, excluding cardiac arrest, requiring a patient to be seen by a critical care outreach team (CCOT) due to vasovagal attack, breathing difficulty, bleeding, loss of consciousness, seizure, cardiac tamponade, chest pain or pre-arrest rhythm.

### Data processing

Data were extracted for patients admitted from January to December 2020. Patients’ demographics, physiological parameters, NEWS2 score, death and transfer to ICU were extracted from EHR. Diagnosis and comorbidities were gathered using ICD-10 coded data. Cardiac arrest and medical emergency were extracted from the CRT database. Patients with positive COVID-19 cases were identified from the COVID-19 pathology data, a daily updated database submitted to NHS England. Data for critical events and COVID-19 cases were linked to extracted EHR data using SQL by clinical data analysts (led by NK) in the hospital. Data was Pseudonymised, transferred to the principal investigator (BA) via the NHS network, and then moved to UCL data safe haven (DSH) for data analysis. The DSH is a secured database system with restricted access to the PI and research team, via safe gateway technology.

### Statistical analysis

Analysis was done in the DSH using the R programme. Data cleaning and pre-processing were done by BA. Dependant and independent values, categorical and missing values, and splitting data into training and test data sets were all addressed prior to analysis.

Statistical significance was defined as a *p*-value of <0.5 using two-tailed tests.

Assessment of missing data in NEWS2 scores was done using *t*-test to compare with another complete variable to identify association with other variables or random missingness.

The categorical variables are presented as percentages (count), and the continuous variables are presented as the mean ± standard deviation. The normality analysis of the data was assessed using Box plots for the frequencies. The inter-group difference between categorical variables was evaluated using Pearson’s chi-squared test. The difference between groups was compared using the Mann–Whitney U test for non-normally distributed data.

The correlations between NEWS2 and physiological parameters and outcomes were evaluated using the Pearson correlation coefficient. Correlation coefficient values range between -1.0 and 1.0, where -1.0 shows perfect negative correlation and 1.0 indicates perfect positive correlation. To supplement the model with parameters that could improve the prediction, we split data into training and testing datasets using the Train/Test method (70% for training and 30% for testing). Univariate and multivariate logistic regression analysis was conducted to assess the association between score and outcomes. The prognostic value of NEWS2 and supplemented model for hospital death, transfer to ICU, cardiac arrest and medical emergency were evaluated using the receiver operating characteristic (ROC) analysis. The value of the areas under the ROC curve (AUC) was measured. The cut-off points of the models were assessed using Youden’s index: sensitivity, specificity, positive and negative predictive values. The AUC’s values were interpreted using reported criteria by Fischer et al.: AUC > 0.9, 0.7 to 0.9, and 0.5 to 0.7 indicate high, moderate and low predictive accuracy, respectively (17).

## Results

### Baseline characteristics

The initial cohort comprised of 16978 admitted patients, forming 40901 encounters and 68867 admissions to the wards from various specialities in oncology, cardiology, medicine and surgery. Patients with a primary cardiovascular disease diagnosis were 7313 (36%), 14798 encounters and 24792 ward admissions. Patients with missing NEWS2 or physiological parameters values were 21% of the total cohort and 16% of cardiac patients. Using the *t*-test, the means of NEWS2 and age were similar (60.3 and 59.8, respectively) with a statistical significance of *p-*value < 0.01. Therefore, considered Missing completely at random (MCAR)(18) and can removed (19). Included cardiac patients were 6143 patients admitted under cardiology, cardiothoracic surgery, congenital heart diseases, or cardiac surgery specialities (Appendices 1). Patients with COVID-19 were 248 (4%), 40% were cardiac patients.

The mean age of the cardiac population is 63.73+14.47, and 69% were males. The in-hospital mortality was 12% (743 patients), ICU admission was 15% (921 patients), and 117 cardiac arrests and 160 medical emergencies. The characteristics of the study population are tabulated in table 1. The difference between dead cases, patients admitted to ICU, who developed cardiac arrest, or medical emergencies, and those who did not develop critical outcomes according to the NEWS2 scoring category were statistically significant (p < 0.001). The comparison is tabulated in table 2.

**Table 1.** General Characteristics of patients

| **Variables** | **Descriptive (n=6143)** |
| --- | --- |
| Age, years | 63.7 ± 14.47 |
| *Gender* |  |
| Male | 4238(69%) |
| Female | 1904(31%) |
| *Speciality* |  |
| Cardiology | 3808 (62%) |
| Cardiothoracic surgery | 2027 (33%) |
| Congenital heart disease | 184 (3%) |
| Cardiac surgery | 122 (2%) |
| *Covid patients* | 248 (4%) |
| Cardiology and cardiothoracic surgery | 100 (40%) |
| Other (Oncology, Haematology, Respiratory and Thoracic surgery) | 148 (60%) |
| *NEWS2 numerical parameters* |  |
| Systolic blood pressure, mm Hg | 128.3±23.2 |
| Mean arterial pressure, mm Hg | 91.21±14.1 |
| Pulse rate, beats/min | 74.43±16.7 |
| Temperature, °C | 36.5±0.6 |
| Oxygen saturation, % | 96.7±2.23 |
| Diastolic blood pressure, mm Hg | 72.7±11.9 |
| NEWS2 | 1.5±1.7 |
| *Outcomes* |  |
| In hospital mortality | 743 (12%) |
| ICU admission | 921 (15%) |
| Cardiac arrest | 117 (2%) |
| Medical emergency | 160 (3%) |

**Table 2.** Comparison between categorical parameters of study population

|  | NEWS 2 categories |  |  | P value |
| --- | --- | --- | --- | --- |
| characteristic | Low (0-4) | Moderate (5-6) | High (7=<) |  |
| *Speciality* |  |  |  |  |
| Cardiology | 3431 | 249 | 128 | >0.001 |
| Cardiothoracic surgery | 1931 | 86 | 10 | >0.001 |
| Congenital heart disease | 164 | 17 | 3 | >0.001 |
| Cardiac surgery | 113 | 6 | 3 | >0.001 |
| *Outcomes* |  |  |  |  |
| In hospital mortality | 597 | 62 | 84 | >0.001 |
| ICU admission | 850 | 36 | 35 | 0.013 |
| Cardiac arrest | 85 | 14 | 18 | >0.001 |
| Medical emergency | 140 | 12 | 8 | >0.001 |

The mean of NEWS2 in death cases was higher by a small difference than alive patients (difference = 1.005, 95%CI, p<0.001). Between ICU admission and non-admitted cases, the mean was similar (difference = 0.01, 95% CI, p<0.09). Between cardiac arrests and non-arrest cases, and medical emergency and stable cases, there was a small variation (difference= 1.99, 95%CI, p<0.001 & difference= 0.99, 95% CI, P<0.001, respectively) (Figure 1,2,3 &4).

**Figure 1.**
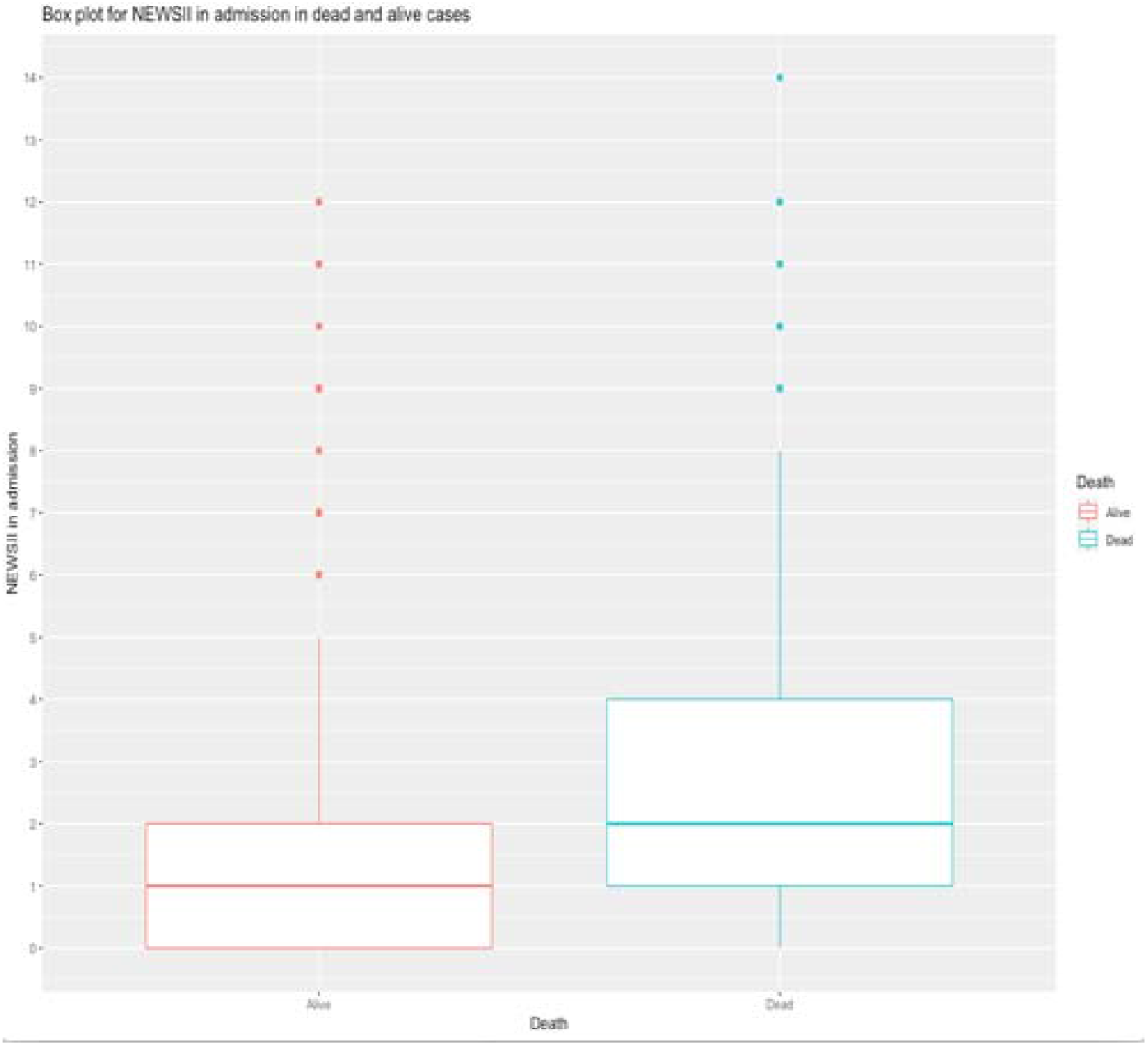
Box plot for NEWS2 in survived and non-survived cases.

**Figure 2.**
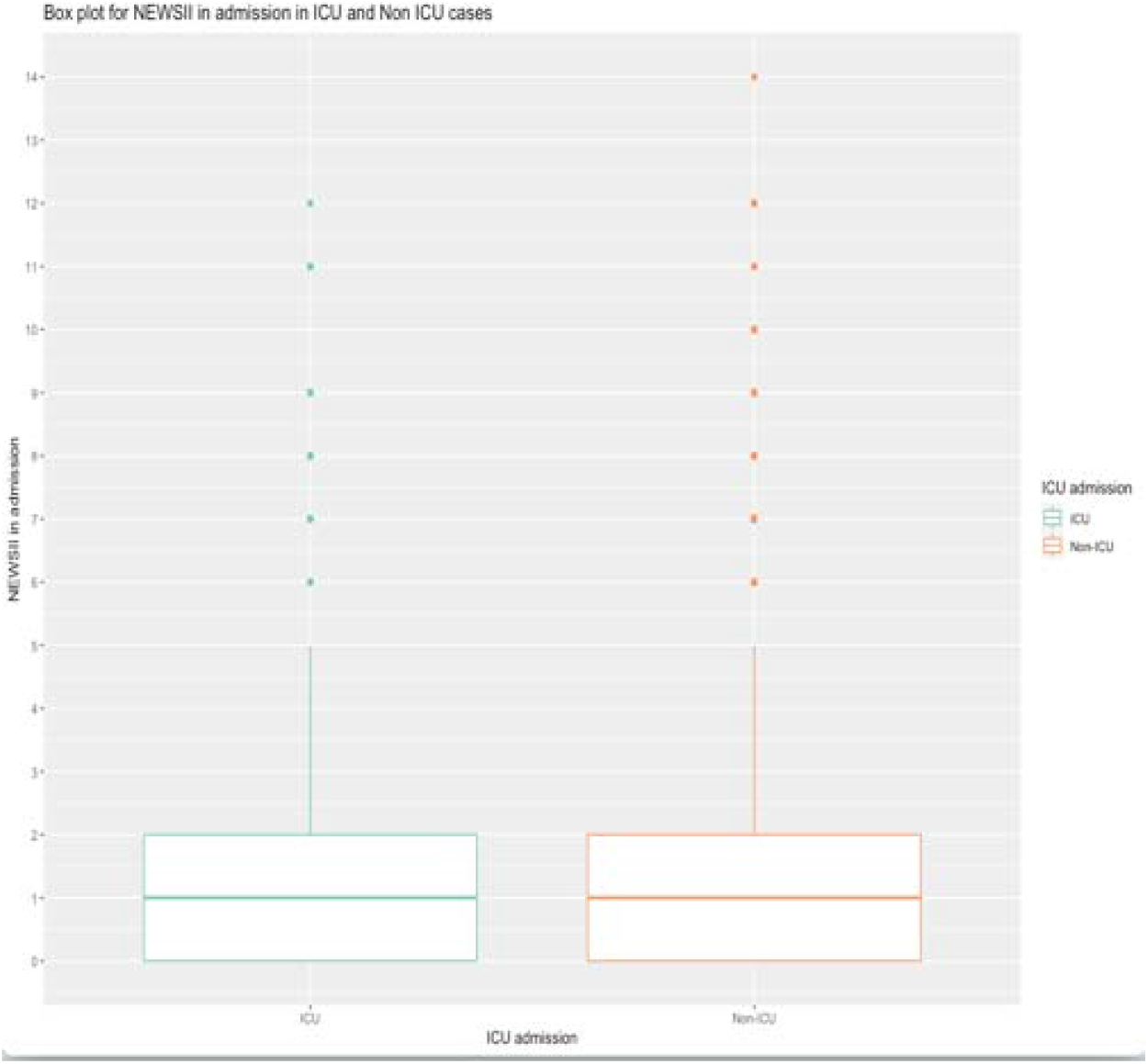
Box plot for NEWS2 in ICU admission and non-ICU cases.

**Figure 3.**
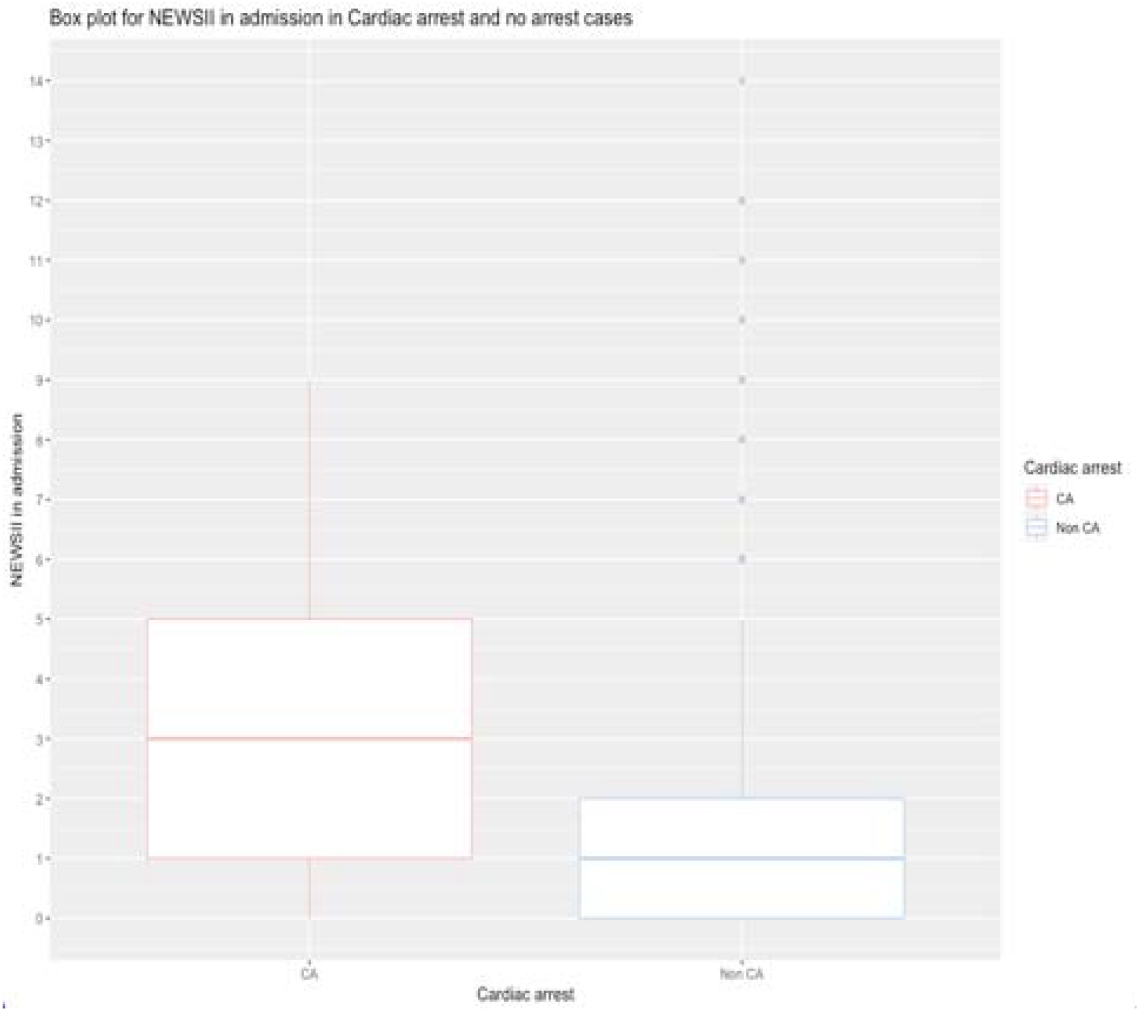
Box plot for NEWS2 in cardiac arrest and non-arrest cases.

**Figure 4.**
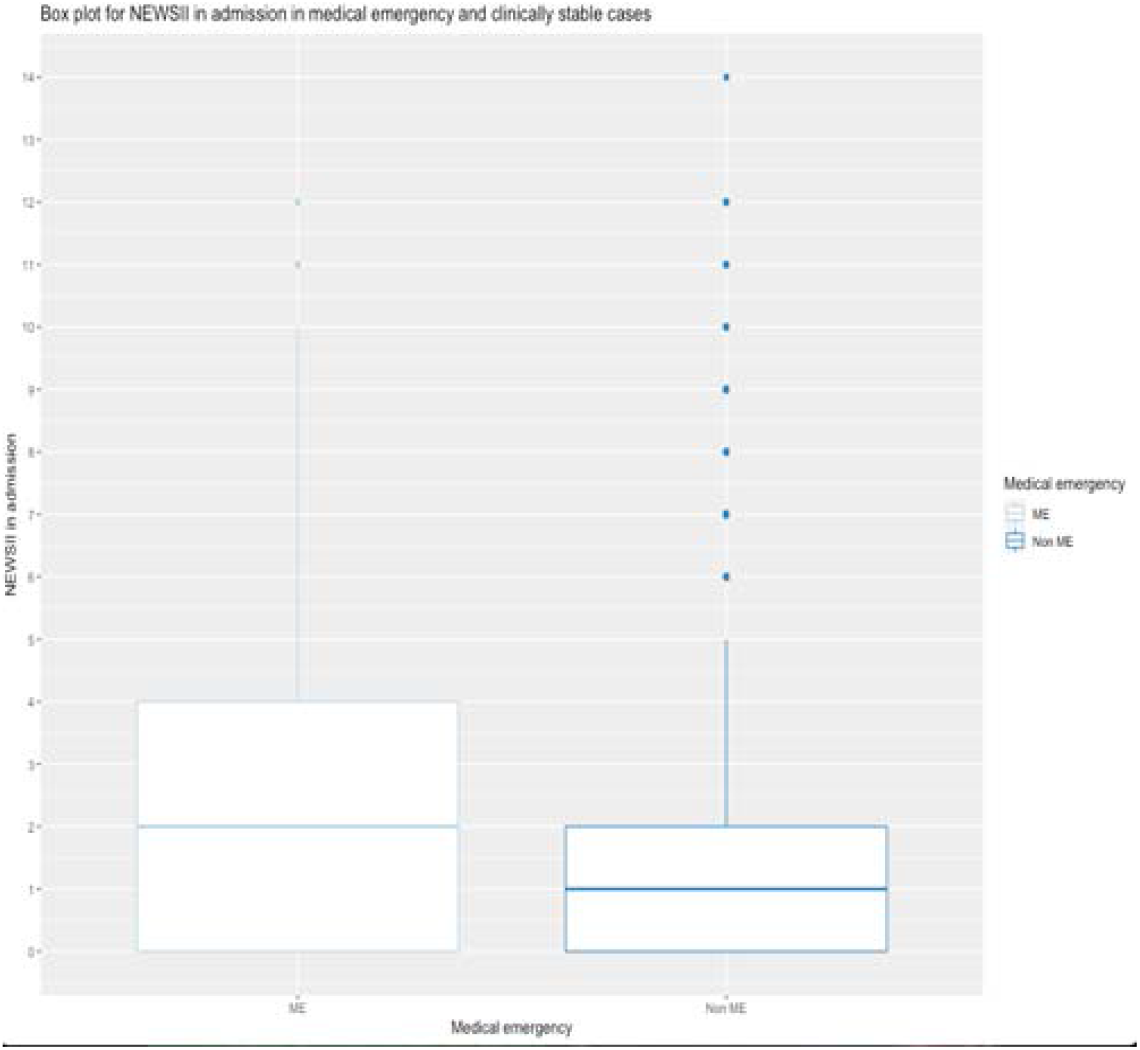
Box plot for NEWS2 in Medical emergency non-emergency cases.

Using the correlation matrix between parameters measured on admission and outcomes, we found a positive correlation between temperature with heart rate (0.32); respiration rate, heart rate and death with NEWS2(0.41,0.31,0.30); and death with cardiac arrest (0.31). In the parameters 24 hours prior to critical events, there was a strong correlation of SpO2 with Systolic pressure, CVPU with NEWS2(0.42,0.41,0.42), and Systolic pressure, SpO2 and death with age (0.30, 0.31,0.34). Cardiac rhythm is strongly correlated with cardiac arrest and death (0.51,0.90) (Figures 5 &6).

**Figure 5.**
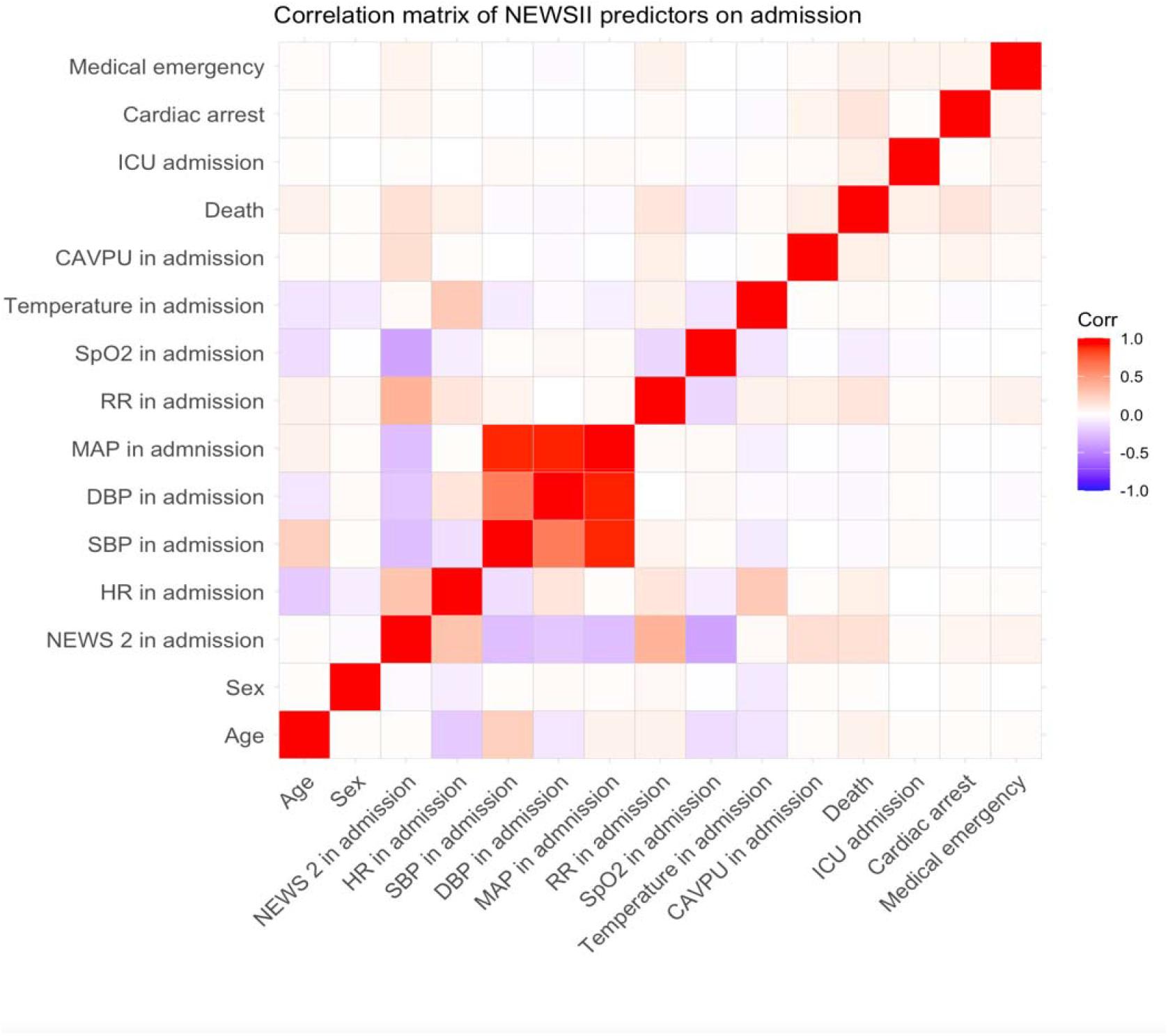
Correlation matrix using Pearson’s correlation coefficient between parameters and NEWS2 on admission and outcomes.

**Figure 6.**
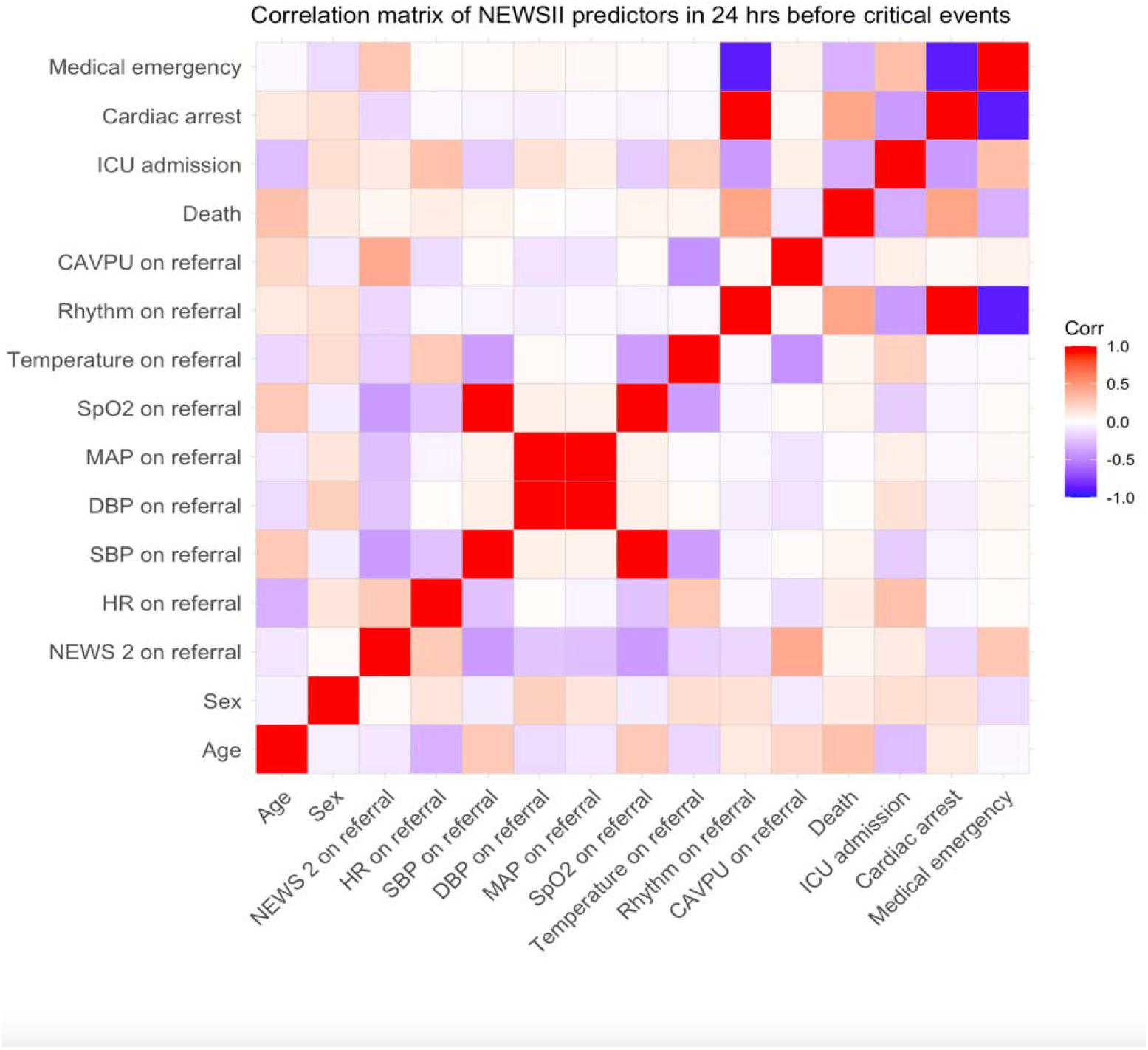
Correlation matrix using Pearson’s correlation coefficient for parameters and NEWS2 24 hrs before outcomes.

Regarding the discrimination, NEWS2 showed moderate to low predictive accuracy with death, ICU admission, cardiac arrest, and medical emergency (AUC: 0.63, 0.56, 0.7& 0.63; 95% CI) respectively, while NEWS2 24 before event showed low predictive value (AUC: 0.57, 0.61, 0.53 & 0.56; 95% CI) respectively. In patients with Covid-19, NEWS2 showed good to poor performance (AUC: 0.64, 0.5, 0.81& 0.81; 95%CI). When NEWS2 was supplemented with age, there was insignificant change in the predictive performance for all patients (AUC:0.63, 0.5, 0.73 & 0.64; 95% CI) However, there was significant improvement for Covid patients (AUC: 0.96, 0.7, 0.87& 0.88; 95% CI). This was also true for the model of NEWS2 supplemented with heart rhythm for the cardiac patients (AUC: 0.75, 0.84, 0.95 & 0.94; 95%CI). The calculated optimum cut off value for NEWS2 was ≥5 showed sensitivity for NEWS2 model of 20% and specificity of 94%, while for the model supplemented with heart rhythm sensitivity was 30% and specificity 85% (Table 3 & appendices 2).

**Table 3.** NEWS2 as a predictor of critical events compared to supplemented NEWS2.

| Model | Death<br>(AUC 95% CI) | ICU admission<br>(AUC 95% CI) | Cardiac arrest.<br>(AUC 95% CI) | Medical emergency<br>(AUC 95% CI) |
| --- | --- | --- | --- | --- |
| *Cardiac patients* |  |  |  |  |
| NEWS2<br>(admission) | 0.63<br>(0.58-0.67) | 0.55<br>(0.51-0.62) | 0.69<br>(0.65-0.74) | 0.62<br>(0.57-0.67) |
| NEWS2<br>(24 hrs before<br>outcome) | 0.58<br>(0.53-0.65) | 0.63<br>(0.58-0.69) | 0.54<br>(0.51-0.67) | 0.56<br>(0.52-0.60) |
| NEW2<br>+Age | 0.63<br>(0.58-0.69) | 0.53<br>(0.50-0.59) | 0.73<br>(0.68-0.79) | 0.64<br>(0.59-0.68) |
| NEWS2<br>+Rhythm | 0.75<br>(0.67-0.80) | 0.84<br>(0.78-0.88) | 0.95<br>(0.89-0.98) | 0.94<br>(0.89-0.97) |
| *Covid patients* |  |  |  |  |
| NEWS2 | 0.64<br>(0.59-0.69) | 0.51<br>(0.50-0.56) | 0.81<br>(0.75-0.86) | 0.80<br>(0.74-0.86) |
| NEWS2<br>+Age | 0.96<br>(0.89-0.98) | 0.69<br>(0.63-0.76) | 0.87<br>(0.81-0.94) | 0.88<br>(0.82-0.94) |

## Discussion

Our retrospective study is among the first to evaluate the prognostic power of the digital early warning score NEWS2 in a patient with CVD in a specialist cardiac setting. We were able to validate NEWS2 in patients with COVID-19 in the cardiac setting. The main findings of the study reveal that: (1) NEWS2 is inadequate on its own to predict deterioration in patients with CVD in the examined specialist cardiac setting; (2) adjustment of the tool by supplementing with positively correlated parameters can strengthen the prognostic performance and therefore reduce the burden of critical events associated with CVD.

Our findings were consistent with a previous study in patients with chest pain (18) while contrary to findings by a study that examined a subset of CVD patients in a single hospital(15). MEWS showed low predictive accuracy in patients with chest pain in ED (18) However, good discrimination was found of ViEWS in a subset of patients in Canada a decade ago and using a distinct EWS than the current NEWS2 (15). Studies were limited in number, scope and population and varied in EWS models. Patients with “normal” vital signs may be sicker than they look through traditional routine monitoring (19). When EWS was explicitly developed for post-cardiac surgery patients, prognostic accuracy was excellent in predicting ICU mortality (20). It included a range of organ system-specific parameters that correlate with cardiac surgery outcomes, such as lactic acid, FiO2 and platelets. Relevant parameters were found using machine learning, including clinical signs and heart rate variability, to improve scoring systems for adverse cardiac events (18). The cardiac rhythm combined with NEWS2 in our study, and the heart rate variability, such as the average of the instantaneous heart rate (avHR) or Ratio of LF power to HF power (LF/HF) selected by Nan et al. (2014), may not be routinely measured or readily available for clinicians as SBP or temperature. Yet, their predictive value indicates the need for highly illness-correlated parameters to be present for clinicians through facilitating timely and thorough assessment.

The subset of patients admitted with Covid-19 showed improvement in NEWS2 when the model was adjusted with age. The finding is supported by a study that reported low to moderate discrimination of NEWS2 for the severity of COVID-19 disease(21). Adjusting the model with age alone did not improve prediction in the study by Ewan et al. (2021), as shown in our findings. However, supplementing with age, routinely collected blood and physiological parameters enhanced discrimination in a multisite (UK and non-UK hospitals) study(9), which seems to further support our results yet may detect the need for additional criteria adjustment. Our study indicates that EWS are not a stand-alone rapid assessment tool as it accords with reported limitations in rapid risk scores for predicting cardiovascular complications (22,23). In clinical settings, and before implementing EWS, clinicians look for signs of abnormality related to body organs affected, disease pathophysiology, or procedure side effect to critically assess the situation. Systems that existed to stratify the risk of long term cardiac complications have been successfully validated and utilised for years, such as the thrombolysis in myocardial infarction score (TIMI) (24) and GRACE (25).

They included cardiac disease variables: heart rate variabilities and serum cardiac biomarkers, which may not be available routinely or at the first admission presentation for rapid assessment. It was also observed that combining nurses’ objective assessment with traditional EWS in the Dutch-Early-Nurse-Worry-Indicator-Score (DENWIS) improved the prediction of ICU admission and mortality in surgical patients (26). Therefore, thorough tracking of short-term deterioration parameters to develop decisive intelligent scoring systems will potentially outperform EWS in various diseases and settings.

It is essential to consider possible issues in developing EWS for specialised subgroups. A potential complexity may be present when having a variety of multiple parameters measured at various times during admission to form a scoring system that is meant to be simplistic and standardised. In addition, the endpoints favoured by researchers in validation studies may not be the ideal points to measure triggered EWS against. In the clinical application of EWS, a high score triggers an action to prevent a critical event (27). In the event of clinically intervening at the right time, the examined adverse events may not occur. Prior to reaching or while preventing an adverse event, precise and proper deterioration endpoints may be more fitted than traditionally studied outcomes, such as death and ICU admission. At the current time of available EHRs integration and data science techniques, it is possible and may be more valid to identify and define appropriate critical illness endpoints to examine EWS against.

We assessed the performance of digitally integrated EWS; the integration generates NEWS2 in patients’ charts from remotely captured parameters by automated monitoring. EHR integration and automation can improve the accuracy and alerting promptness(28). We were able to extract a good sample of CVD patients and identify patients diagnosed with Covid-19 despite the missingness of some NEWS2 recordings. The issue in recording completion could be due to a lack of staff adherence to routine and timely monitoring, as each measurement would be automatically transmitted to patients’ charts.

Therefore, careful and selective modelling of algorithms from parameters that can be available routinely and reflect significant clinical meaning is needed. The validity of NEWS2 in specialist settings like cardiology indicates the need for either score enhancement or systemic supplementing and finer endpoints definition for better detection. Studying the clinical environment from a practical side of EWS will explain the adoption and implementation role in the success or failure of EWS. From various specialities, clinicians’ involvement in models’ development and validation is invaluable to produce a higher accuracy and finely clinical expertise-born warning scores.

## Strengths and limitations

Our study is the first to examine the performance of universal EWS (NEWS2) in patients with cardiovascular diseases in a cardiac specialist hospital. We were able to extract data from EHRs systems where NEWS2 is integrated and automated, reflecting the accuracy of captured parameters and enabled us to integrate other data sources for critical outcomes and COVID-19 cases. The study followed a retrospective data collection from three data sources where there was less control of missingness of NEWS2 recordings, heart rhythm at several points in time, and other parameters that could be examined like Fio2 level. We conducted external validation of NEWS2 and internal of the supplemented model; external validation studies are needed for generalisability.

## Conclusion

The early warning score (NEWS2) in patients with cardiovascular diseases is suboptimal to predict deterioration early. Adjusting early warning scores with variables that strongly correlate with critical cardiovascular outcomes will improve the early scoring models. Thorough tracking of parameters in EHRs and data availability can support the generation of decisive, intelligent models for a readily feasible system in routine clinical work. There is a need for defining and revising critical endpoints and the involvement of clinicians in models’ development that reflect a significant meaning for deterioration detection. Further validation and implementation studies in cardiac specialist settings and other specialist subgroups are required to investigate methods needed to enhance the capacity of EWS where it is least investigated.

## Supporting information

STARD checklist

## Data Availability

Data sharing statement: All data relevant to the study are included in the article or uploaded as supplemental information.

## Other information

### Ethics statement

The study is approved by HRA and HCRA, REC reference: 20/PR/0286

### Patient consent for publication

Not applicable.

### Contributors

AB, DM and TB conceived the study. BA carried the data collection with guidance of DM. BA conducted analysis and interpretation of findings. BA wrote the manuscript, and all authors contributed to the interpretation and revision of the manuscript.

### Funding

BA has received PhD funding from the Saudi Arabian Cultural Bureau to conduct the study. Grant number: ELP003964.

### Competing interest

No competing interest declared.

### Data sharing statement

All data relevant to the study are included in the article or uploaded as supplemental information.

### Twitter

@BaneenAlhmoud, @DanBartsICU, @amibanerjee1, @TimBonnici, @DrRiyazPatel.

## Notes

### Competing Interest Statement

The authors have declared no competing interest.

### Author Declarations

This study is registered and approved by the Health Research Authority (HRA) and Health and Care Research Wales (HCRW) and sponsored by University College London (UCL). REC reference: 20/PR/0286. A confirmation of capacity has been received from St Bartholomews hospital.

