## Appendices 1 and 2 for "Performance of digital Early Warning Score (NEWS2) in a cardiac specialist setting: retrospective cohort study"

1- Flowchart of patients’ cohort and data sources

Included cohort n: 6391 (6134 cardiac & 248 COVID-19 patients)

Non cardiac cases n: 9665

Missing NEWS2. n: 1170

COVID-19 pathology: Positive COVID-19: 248

Cardiac cohort n: 6134

Cardiac cohort n: 7313

EHR: Death and ICU admission: 4912

CRT: Cardiac arrest and medical emergency: 338

EHR: Full cohort n: 16978

Abbreviations: EHR: electronic health record, CRT: cardiac resuscitation team, n: number of patients.

2- Predictive ability of NEWS2.
A. Predictive ability of NEWS2 on admission for death, ICU admission, cardiac arrest, and medical emergency.
1- Death	AUC:0.63	2. ICU. AUC:0.56

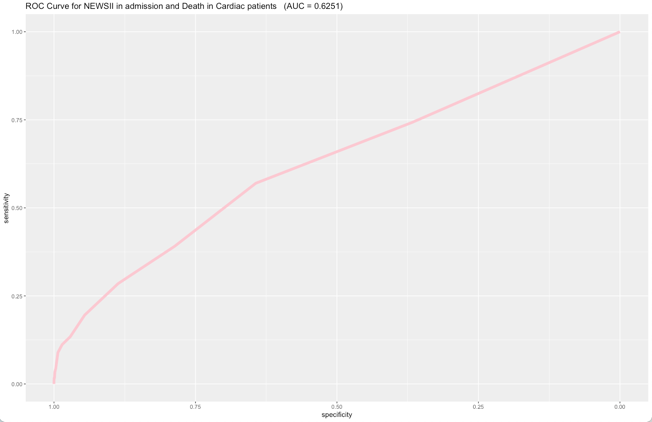

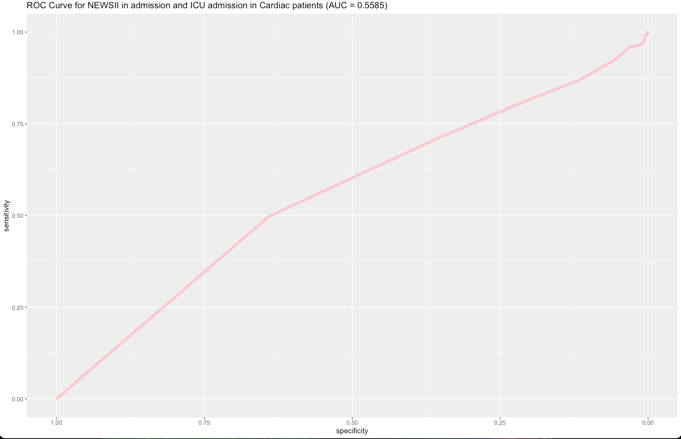

3. CA	AUC:0.69	4. ME AUC: 0.63

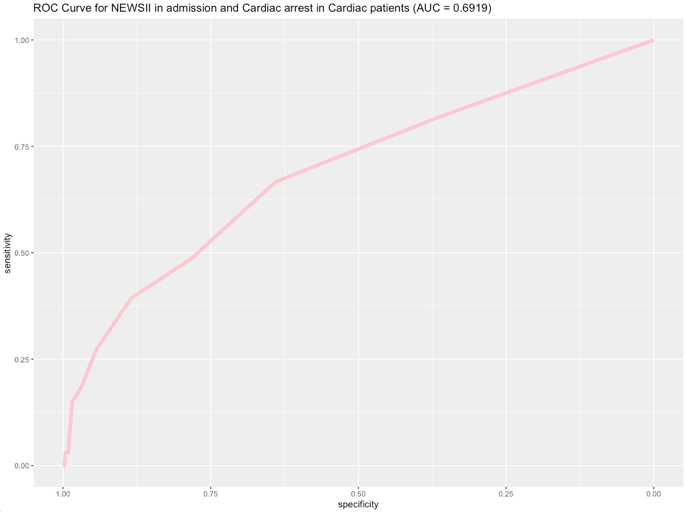

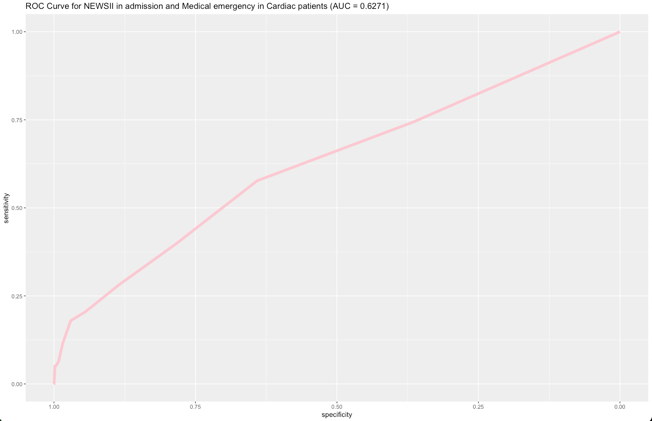

B. NEWS2 predictive ability 24 hours before critical events
1. Death	AUC:0.57	2. ICU. AUC:0.61

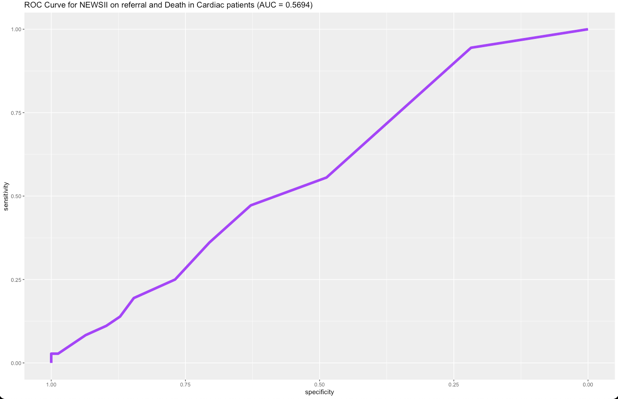

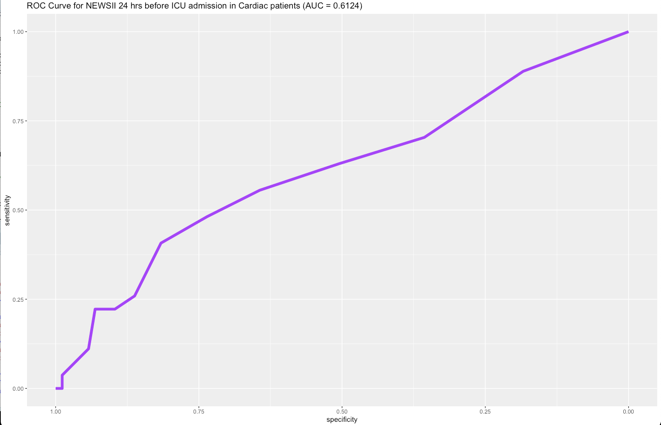

3. CA. AUC:0.53	4. ME. AUC:0.56

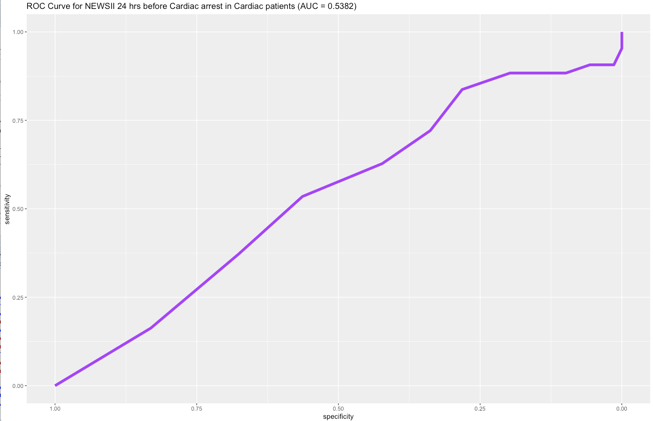

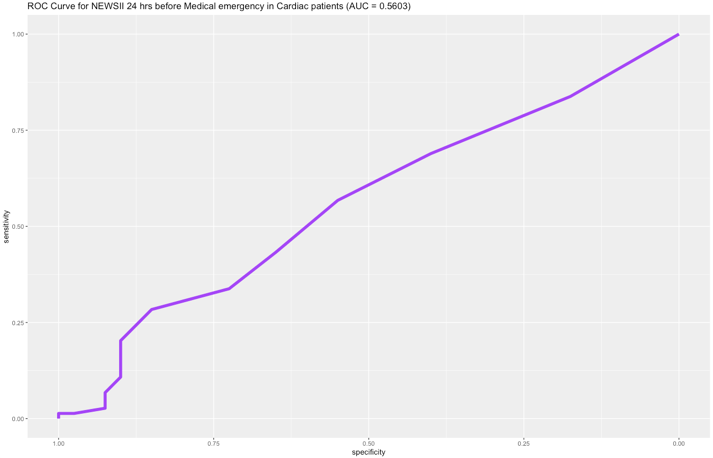

C. NEWS2 predictive ability on admission in patients with COVID-19
1- Death	AUC:0.64	2. ICU AUC:0.5

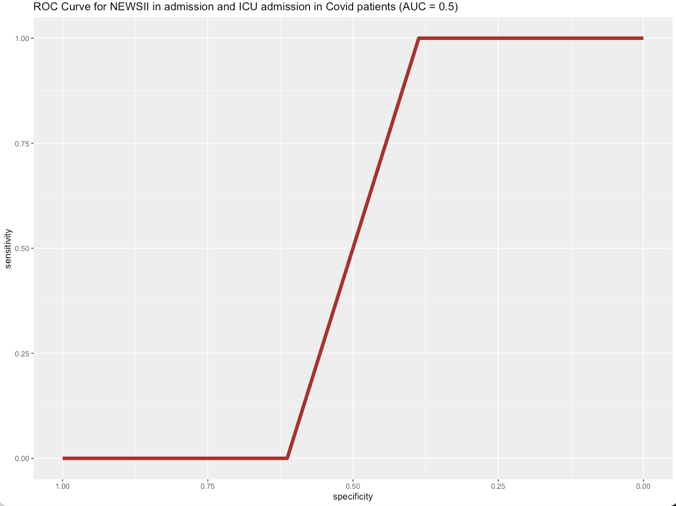

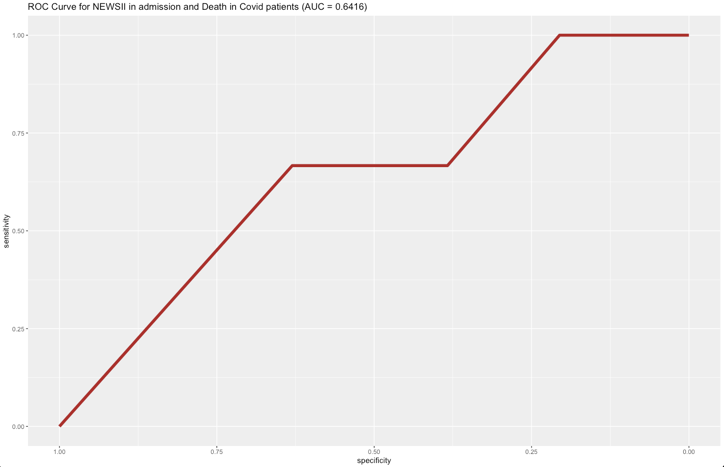

3.CA	AUC:0.81	4. ME. AUC:0.81

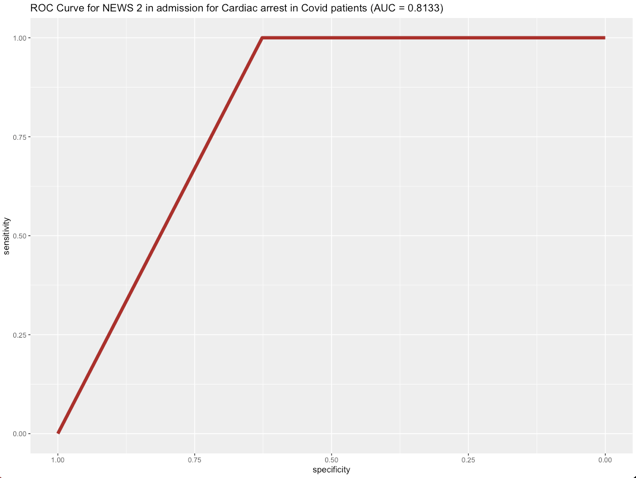

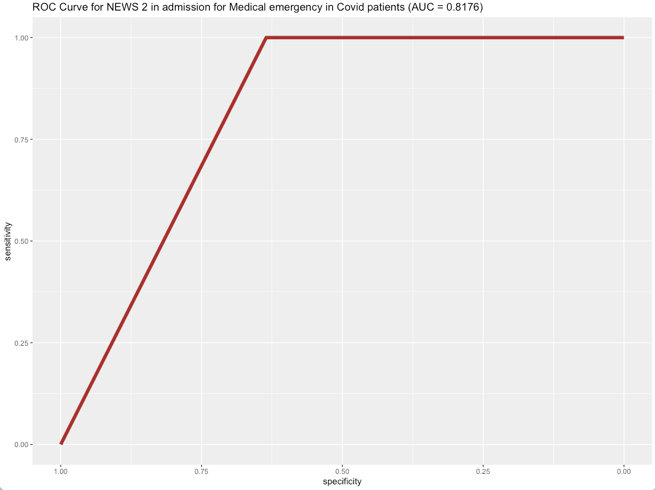

F. NEWS2+age predictive ability in cardiac patients

1- Death	AUC: 0.63	2. ICU	AUC:0.5

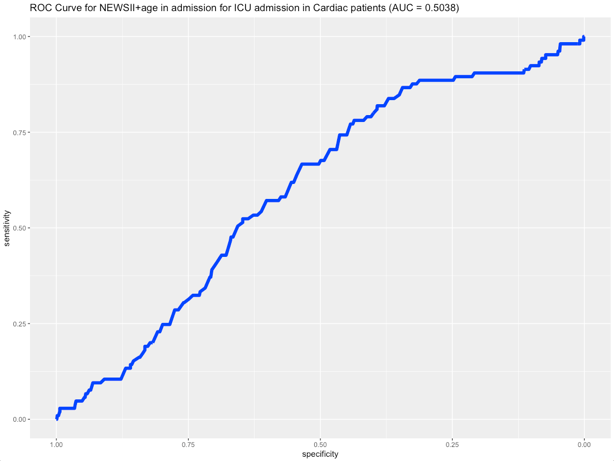

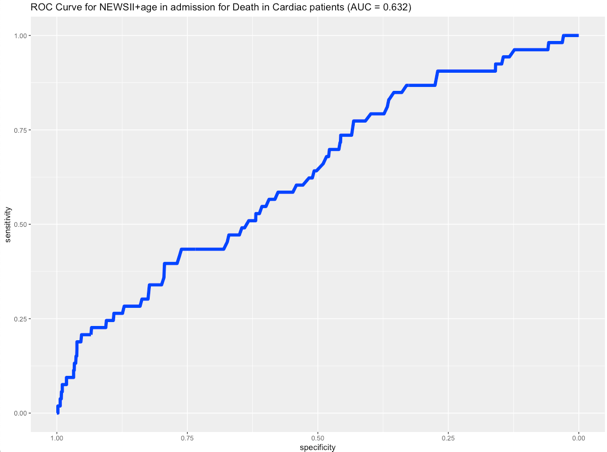

3.CA	AUC:0.73	4.ME. AUC:0.64

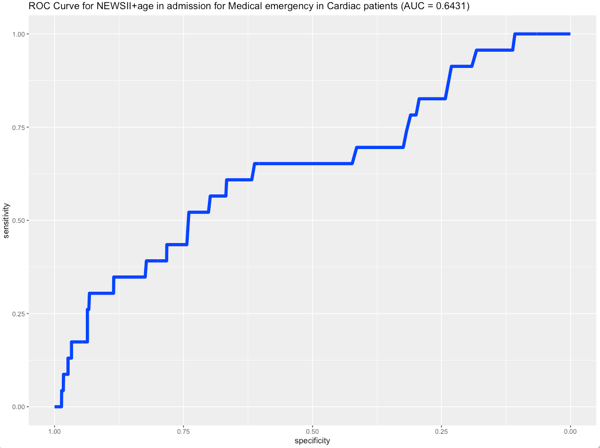

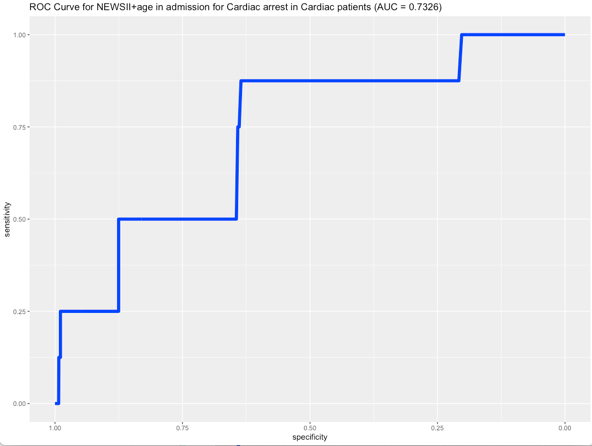

G. NEWS2+ age predictive ability in COVID-19 patients

1.Death	AUC:0.96	2. ICU	AUC: 0.7

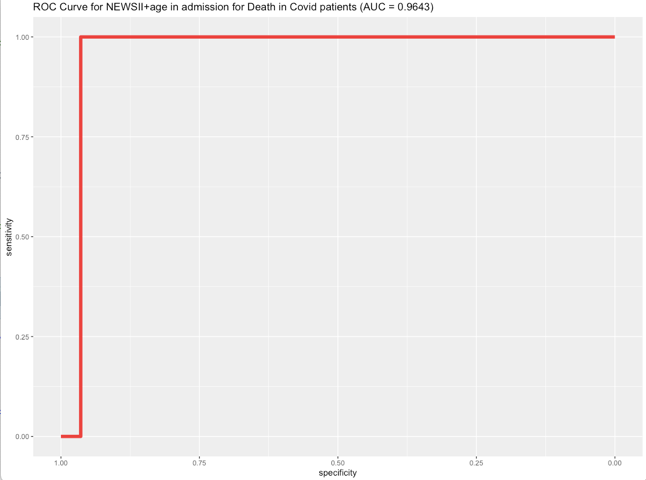

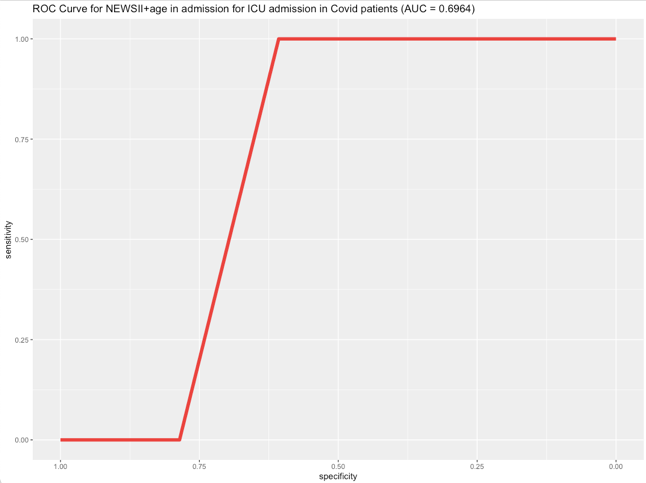

3.CA	AUC:0.87	4.ME	AUC:0.88

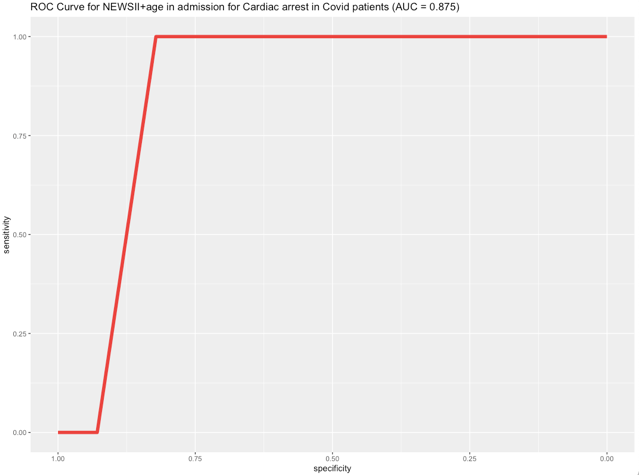

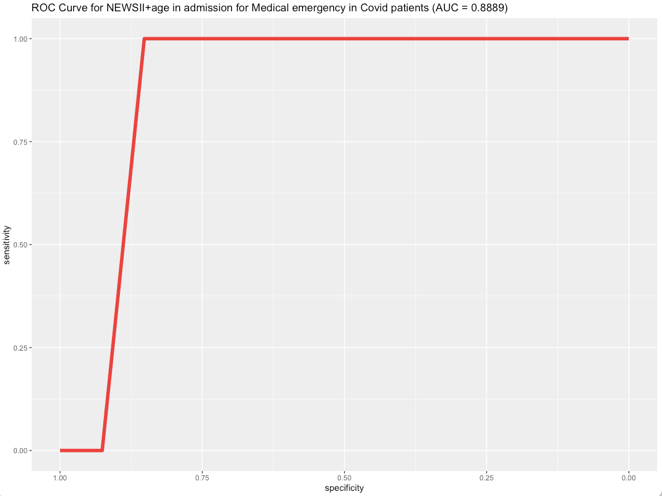

H. NEWS2 +age + cardiac rhythm predictive ability in cardiac patients
1- Death	AUC: 0.75	2. ICU. AUC:0.84

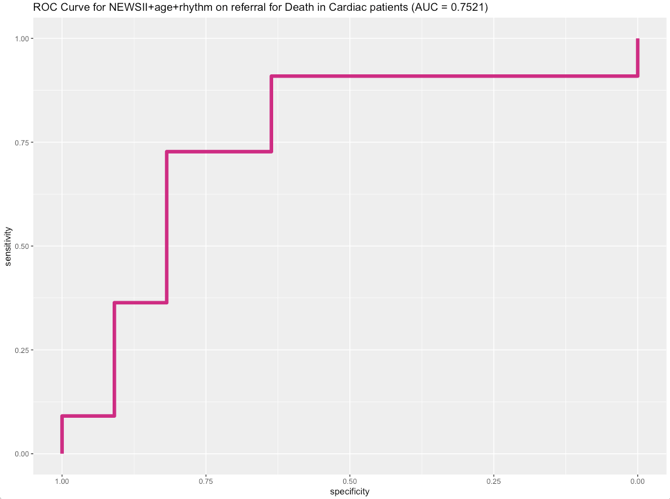

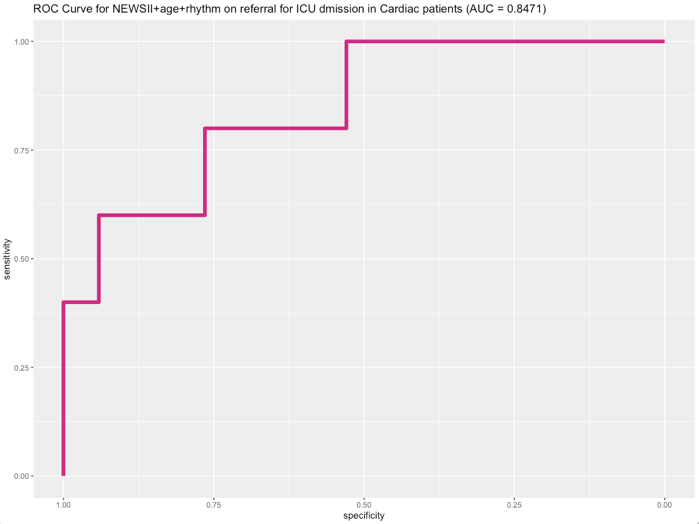

3.CA	AUC: 0.95	4.ME	AUC:0.94

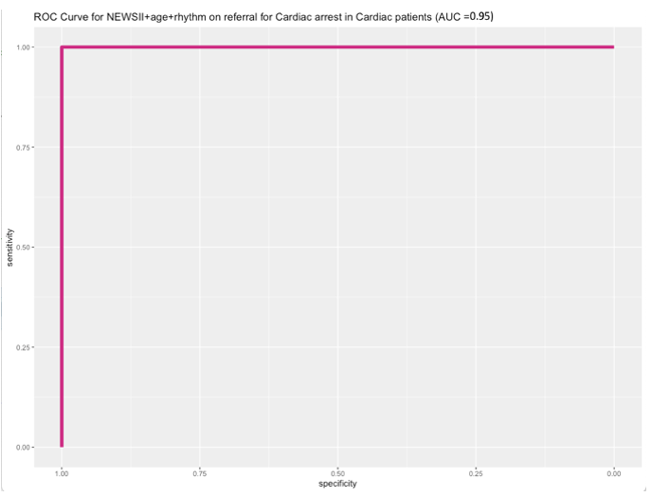

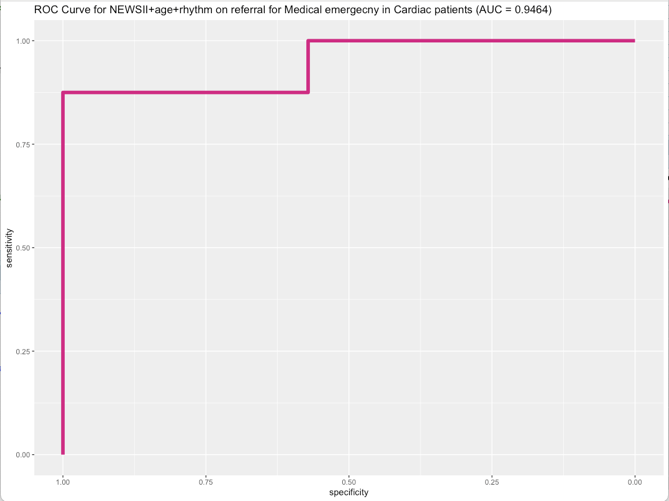

Abbreviations: AUC: are under receiving curve, ICU: admission to intensive care unit, CA: cardiac arrest and ME: medical emergency.
